# Dynamic changes in compressive and shear plantar tissue properties during gait and rest in people living with and without diabetes

**DOI:** 10.64898/2026.03.23.26348696

**Authors:** Chaofan Lin, Athia Haron, Damian Crosby, Garry Massey, Maedeh Mansoubi, Ziyuan Wang, Yufeng Li, Helen Dawes, Andrew Weightman, Glen Cooper

**Affiliations:** School of Engineering, University of Manchester, Manchester, United Kingdom; Medical School, NIHR Exeter BRC, University of Exeter, Exeter, United Kingdom

**Author notes:** Corresponding author: Glen Cooper.

**Keywords:** Mechanical properties of plantar tissue, compressive and shear properties, novel measurement device, ultrasound, gait and recovery, diabetes

## Abstract

Plantar tissue adaptation during activity is thought to contribute to diabetic foot ulceration (DFU), yet most existing studies only measure compressive quasi-static properties.

This pilot study developed an ultrasound-loadcell measurement tool, *PlantarSense*, and used an infrared thermometer to measure dynamic compressive and shear energy dissipation ratio (EDR) and temperature of plantar-tissue at the first metatarsal head (1stMTH) and calcaneus in people living with and without diabetes at baseline, post-walk, and post-recovery.

People living with diabetes showed significantly greater post-walk temperature increases (11.0 % vs 6.9% in controls at calcaneus, p=0.03) and less complete thermal recovery than controls. Baseline compressive EDR at the 1stMTH was significantly higher in people living with diabetes (67.8% vs 56.0% in controls, p=0.04). EDR modulation was greater from shear loading (21.5%) than compression (5.4%) and post-walk induced reductions in EDR were present in all participants, but people living with diabetes showed a 20% lower recovery than controls.

Impaired thermoregulation and tissue adaptation in people living with diabetes was demonstrated by plantar temperature and EDR differences in post-walk and post-recovery. Future work is needed to test more participants with a greater range of diabetes progression to quantify statistically significant plantar tissue differences to inform DFU risk management.

## Introduction

Diabetic foot ulcers (DFUs) affect approximately 19–34% of individuals living with diabetes during their lifetime and are associated with high healthcare costs and serious morbidity [1]. Plantar tissue undergoes repetitive loading during gait, which influences its mechanical integrity and function. In individuals living with diabetes or foot pathology, altered loading patterns and reduced tissue compliance may increase mechanical strain, delay recovery, and elevate ulceration risk. Key functional properties of plantar soft tissue include stiffness and damping, which reduce peak pressures and protect against injury [2]. One useful descriptor of this cushioning function is the energy dissipation ratio (EDR), defined as the proportion of mechanical energy absorbed rather than elastically returned during loading. Changes in EDR reflect altered tissue viscoelasticity and load-bearing behaviour, and elevated EDR has been reported in people with diabetes [3]. Ageing and pathological conditions such as diabetes are associated with structural and mechanical changes in plantar tissue that reduce load-bearing capacity [4] and contribute to pain, falls, and impaired mobility [5]. In people living with diabetes, plantar tissues tend to become stiffer and less compliant, contributing to abnormal loading patterns and increased risk of ulceration [6]. Under loading, plantar soft tissue exhibits a nonlinear stress–strain response, typically characterised by an initial low-stiffness region followed by a stiffer region at higher strain [7]. In addition to normal loading, plantar shear stress, defined as forces acting parallel to the skin surface, has emerged as an important contributor to tissue breakdown and ulcer development [8,9]. Foot skin temperature is also a clinically relevant marker, as localised increases in plantar temperature may indicate inflammation, poor EDR or early tissue injury and may therefore support early DFU detection [10].

Existing studies are difficult to interpret owing to methodological differences between testing protocols and the limitations of static or simplified material models. In particular, in vitro approaches may not accurately reflect physiological behaviour during gait, highlighting the need for physiologically relevant in vivo assessment [11–13]. Because plantar soft tissues are viscoelastic, their behaviour is loading-, time- and temperature-dependent, and may change dynamically during activity and recovery. Walking can alter hysteresis-related energy dissipation, while plantar temperature also changes with activity and differs between people living with and without diabetes [14]. These thermal responses are thought to reflect a combination of altered tissue mechanics, microvascular perfusion and local tissue stress or damage[15–17]. Temperature and mechanical loading energy exchange was recently explored by Haron et al [18] where they explained energy exchange mechanisms and showed significant correlation between cumulative mechanical loading and temperature change in plantar tissue. Together, these observations suggest that dynamic assessment of both plantar temperature and energy dissipation ratio (EDR) may provide complementary insight into tissue adaptation and recovery that is not captured by static assessment alone [15,19–21]. Actual foot–ground interactions involve both normal and shear loading, yet despite the recognised role of shear in tissue damage, its quantitative assessment remains limited due to measurement challenges [22–25]. Ultrasound-integrated in vivo systems are particularly promising in this context because they enable simultaneous measurement of tissue deformation and loading under dynamic conditions [26,27].

To the authors knowledge, no existing in vivo system simultaneously quantifies plantar tissue properties under both normal and shear loading. Furthermore, few studies have directly compared plantar tissue mechanics between people living with and without diabetes or examined how these properties change during activity and recovery. Robust and physiologically relevant methods are therefore needed to characterise plantar soft tissue behaviour across populations and loading conditions.

To address these gaps, this study aims to understand the mechanics of human plantar tissue and how these change alongside temperature changes at baseline, after walking and after a recovery period in people living with and without diabetes. To achieve this an in vivo measurement system, *PlantarSense*, was developed that integrates ultrasound plantar tissue imaging and compressive and shear force measurement from load cells enabling the calculation of plantar tissue energy dissipation ratio (EDR) was used in a gait laboratory walking study. We anticipate that our measurements will show:

### Hypothesis 1

Plantar Surface Temperature Change after walking and after recovery will be greater and lower respectively in people living with than without diabetes.

### Hypothesis 2

Plantar Tissue Energy Dissipation will be different in both compressive and shear for people living with than without diabetes and changes will be greater after activity and slower to recover in people living with than without diabetes.

## METHODS

### 2.1 PlantarSense

Tissue Property Measurement System Development

#### 2.1.1 Measurement System Design

The *PlantarSense* system consists of a linear high-resolution B-mode ultrasound probe (L15-6L25N, Clarus EXT-1M, Vilnius, Lithuania) mounted within a custom aluminium frame and supported by three load cells (LCEB-10, OMEGA, Michigan, USA), enabling all compressive and shear forces applied to the probe head to be transmitted directly through the load cells for accurate mechanical measurement, see figure 2.1A.

**Figure 0.1.**
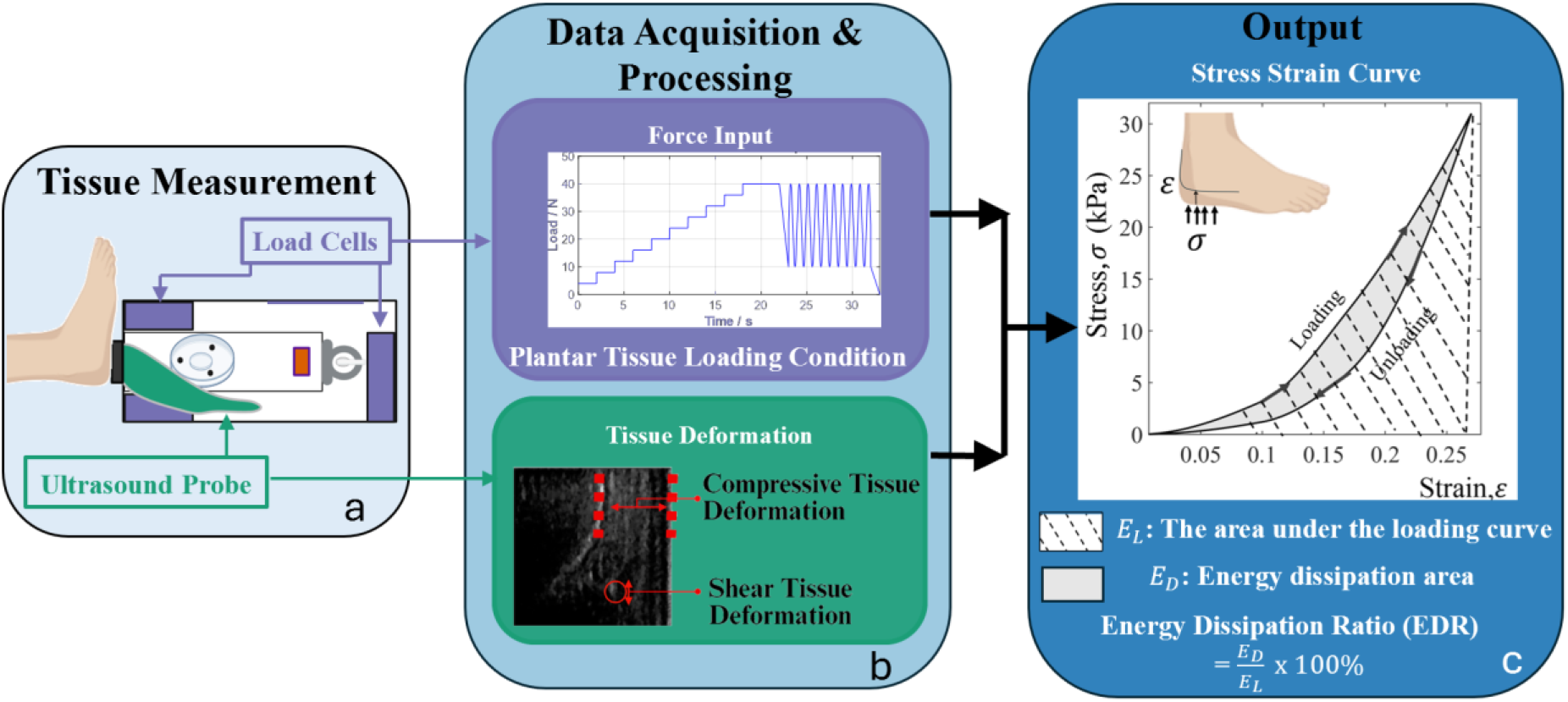
Schematic overview of the plantar tissue measurement system(a), measurement of force and deformation (b) and calculation of energy dissipation ratio (c).

**Figure 0.1.**
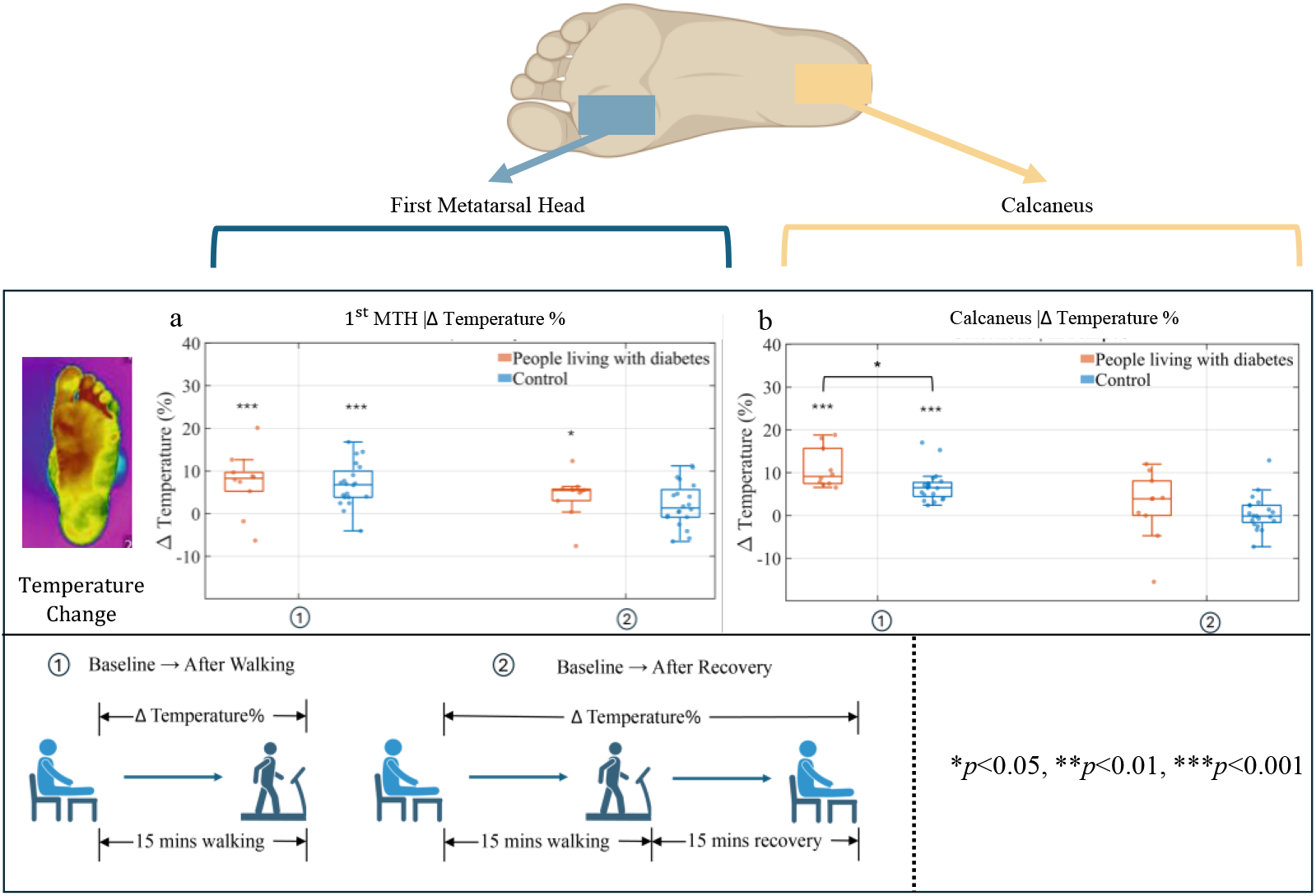
Group comparison of plantar surface temperature changes (ΔTemp%) across phases at the first metatarsal head (1stMTH) and calcaneus. panels (a-b) show ΔTemp% at the 1st MTH and calcaneus, respectively, for people living with diabetes and control groups. Two comparison conditions are presented: (①) Baseline → After Walking, reflecting the effect of 15 minutes of walking on surface temperature; and (②) Baseline → After Recovery, reflecting recovery-related changes following 15 minutes of rest. Unbracketed asterisks indicate within-group tests against zero change (Δ=0), whereas bracketed asterisks indicate between-group comparisons (diabetic group vs control group) at the corresponding timepoint (tests as described in Methods). *p<0.05, **p<0.01, ***p<0.001.

*PlantarSense* measurements included simultaneous acquisition of force and tissue deformation, from which stress–strain curves were generated and material properties were subsequently derived. Force measurement from load cell signals were digitised via a 24-bit high-precision analogue-to-digital amplifier (HX711 ADC, HALJIA, Zhongai, China) and processed in real-time using a microcontroller (myRIO, National Instruments, Texas, USA). All signals were sampled at 60 Hz and visualised via LabVIEW (National Instruments, Texas, US) during acquisition. Tissue deformation measured through ultrasound sequences were captured separately at 60 Hz using EchoWave II software (Telemed, Vilnius, Lithuania). Synchronisation was enabled through a common event (tapping the device 3 times on the foot). Captured data were post-processed in MATLAB (The MathWorks Inc., Natick, MA, USA) using custom scripts to compute stress and strain from *PlantarSense* force and tissue deformation signals, based on the known probe–tissue contact area, and to generate stress– strain curves (workflow shown in Fig. 2.1). A 5 N compressive preload was used for both compression and shear measurements. Energy dissipation was quantified from the loading–unloading cycles. The dissipated energy (*E*_*D*_) is given by equation 1, where *E*_*L*_ is the area under the loading curve and *E*_*U*_ is the area under the unloading curve (see figure 2.1c).

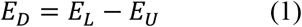

The energy dissipation ratio (EDR, %) is given by equation 2.

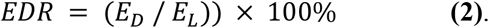

#### 2.1.2 System Validation: Bench Top Mechanical Testing

##### Experimental setup and test method

To address repeatability and accuracy, a silicone phantom (Ecoflex 00-10 silicone, Smooth On Inc., Macungie, USA) and a porcine sample (pork belly with skin and fat layer) were tested, with dimensions of 50 × 50 × 25 mm^3^ and 100 × 100 × 30 mm^**3**^ respectively. Four samples were tested for each loading mode (compression and shear). Each sample was tested both with *PlantarSense* and with a mechanical test machine ((Instron 3344L3928, 100 N load cell, Instron®, Norwood, MA, USA) as illustrated in figure 2.2.

For compression testing, samples were loaded quasi-statically using a 3D-printed indenter replicating the base geometry of the *PlantarSense* ultrasound probe (Fig. 2.2). Quasi-static compression was applied in 2.5 N increments from 2.5 N to 20 N, with each load held for 3 seconds to allow relaxation. Dynamic compression was applied using a sinusoidal waveform (0–20 N) at 1 Hz, corresponding to a loading rate of 45–60 N/s, selected to reflect physiologically relevant plantar stress levels during gait. For shear validation, a custom-designed shear rig (Fig. 2.2) was employed, in which lateral forces were applied under a constant compressive preload of 20 N using the same indenter (adapted from [23]). Quasi-static shear loading was performed in 1 N increments from 1 N to 10 N, each held for 3 seconds, while dynamic shear loading followed a sinusoidal profile (0–10 N) at 1 Hz, corresponding to a loading rate of 20 N/s, simulating shear stresses experienced during walking.

**Figure 0.2.**
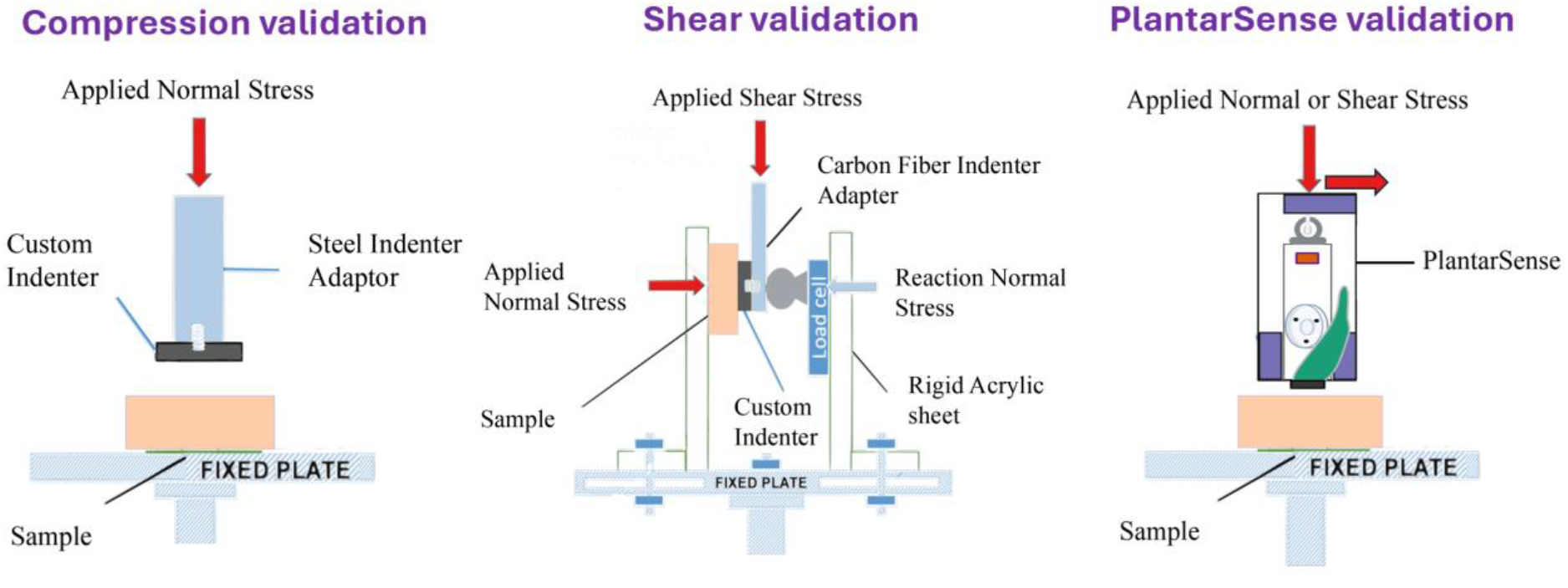
Bench-top validation of custom plantar tissue measurement device under compressive and shear loading conditions (adapted from [23])

**Figure 0.2.**
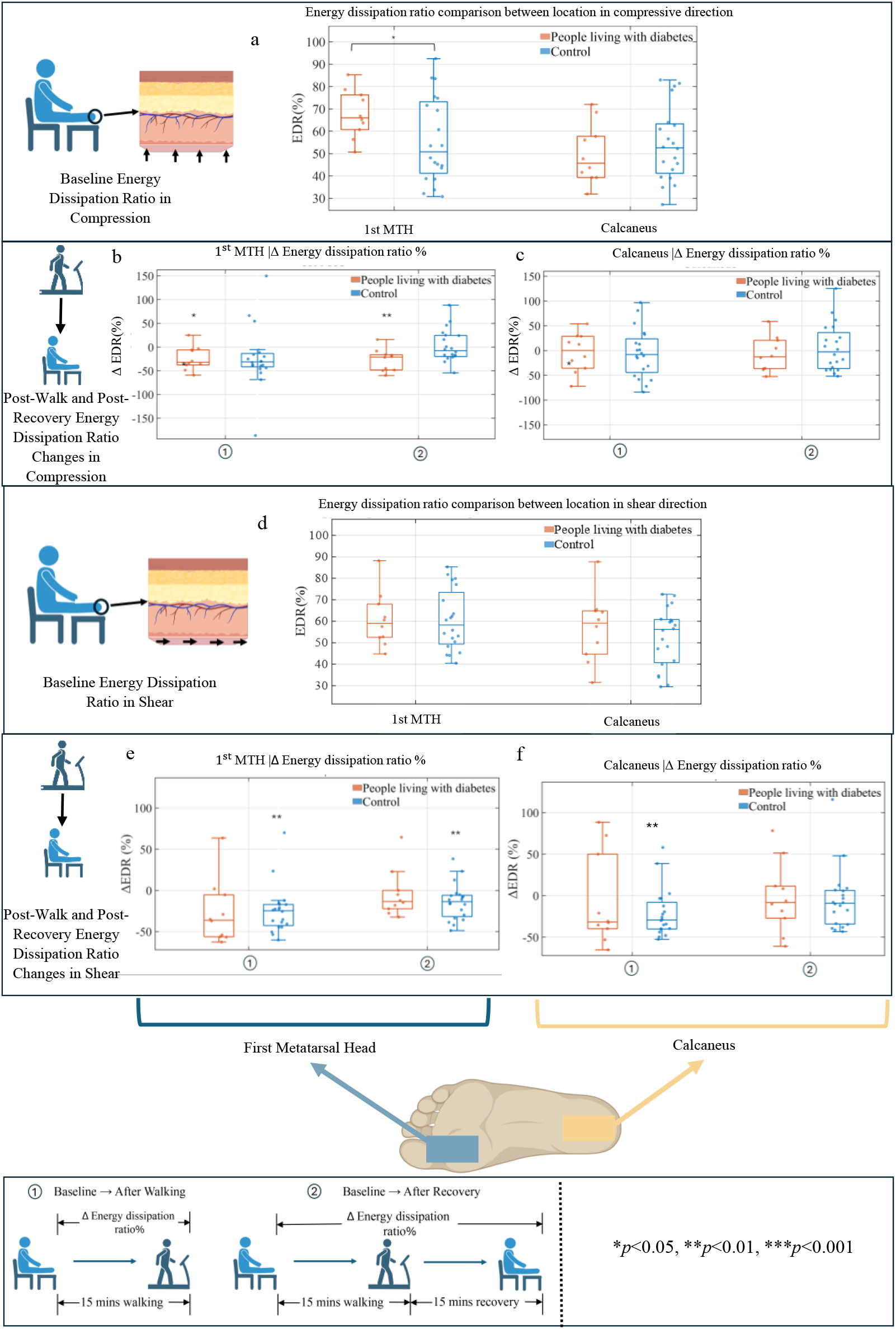
Group comparison of plantar tissue energy dissipation ratio (EDR) at baseline and its activity-related modulation at the first metatarsal head (1st MTH) and calcaneus under compressive and shear loading. Panels show: (a) baseline EDR (%) in compression; (b–c) percentage change in compressive EDR (ΔEDR, %) from ① Baseline → After Walking and ② Baseline → After Recovery at the 1st MTH and calcaneus, respectively; (d) baseline EDR (%) in shear; and (e–f) percentage change in shear EDR (ΔEDR, %) across the same two phase comparisons at the 1st MTP and calcaneus, respectively. Boxplots indicate the median and interquartile range (with individual participant data overlaid).

##### Data Analysis

All data were processed using MATLAB (The MathWorks Inc., Natick, MA, USA), unless otherwise specified. Repeatability of the measurement system was assessed by computing the mean and standard deviation of modulus values across repeated trials on each sample. Coefficient of variation was used as an additional metric for assessing intra-sample consistency. To assess measurement accuracy, modulus values obtained using *PlantarSense* were directly compared to those obtained using the mechanical test machine.

### 2.2 Gait Laboratory Walking and Recovery Plantar Tissue Test in People Living with and without diabetes

Reporting is aligned to the Strengthening the Reporting of Observational studies in Epidemiology (STROBE) reporting guidelines for observational studies and checklist [28].

#### 2.2.1 Ethics and Participant Recruitment

The study received ethical approval from the College of Life and Environmental Sciences, Sport and Health Sciences Research Ethics Committee, University of Exeter (Application No: 22-05-04-B-02 v2), and all participants provided written informed consent prior to participation. All methods were performed in accordance with the relevant guidelines and regulations.

Fifteen participants were recruited for the test: five people living with diabetes (herein referred to as diabetic group) and ten healthy controls without a diagnosis of diabetes (herein referred to as control group). Inclusion criteria required participants to be able to walk unaided for the duration of the experimental protocol and to be free from open wounds, oedema, active ulcers, or severe structural foot deformities. Participants with a history of recent foot surgery or conditions that could affect gait or plantar tissue structure were excluded. The two groups were comparable in demographic characteristics. The mean value (and standard deviation) for the body mass index and age for the diabetic group and the control group was 24.6 ± 3.1 *kg*/*m*^2^, 42.0 ±11.9 years and 23.3 ± 2.7 *kg*/*m*^2^, 40.3 ± 11.5 years respectively, full participant details are in appendix.

#### 2.2.2 Experimental Protocol

The experiment was split into three parts, baseline, walking and recovery illustrated in figure 2.3.

**Figure 0.3.**
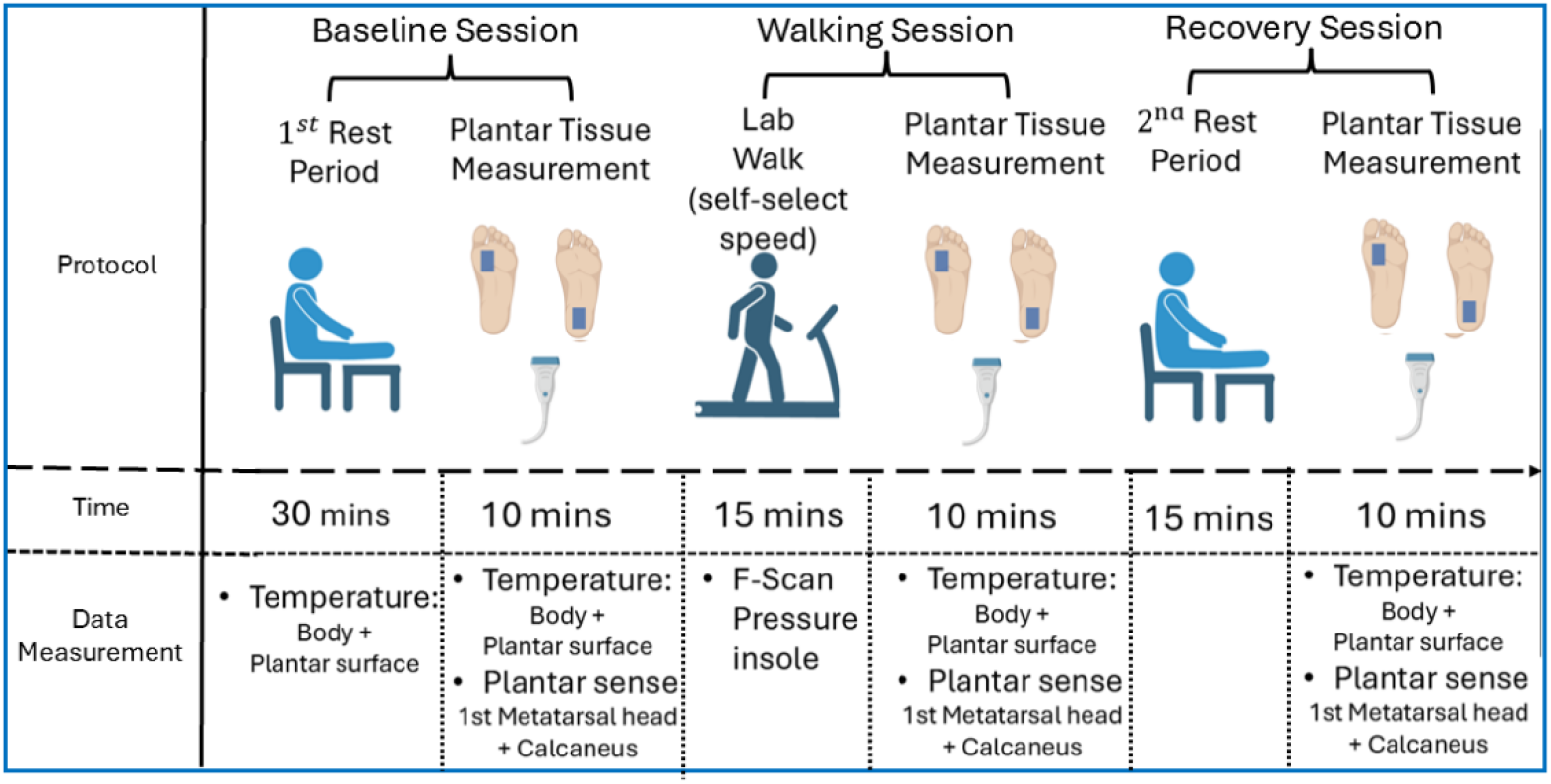
Protocol Overview for gait laboratory walking and recovery plantar tissue test

##### PlantarSense measurement load condition

For compression measurement, quasi-static compression was applied in incremental steps of 2.5N from 0 N to 40 N, with each load held for 3 seconds to ensure tissue relaxation. For dynamic testing, sinusoidal compressive loading (10–40 N) was applied at 1 Hz, corresponding to a loading rate of 45– 60 N/s. Shear measurements were conducted under a constant normal preload of 15 N. Quasi-static shear forces were applied in 1 N increments from 0 N to 10 N, each held for 3 seconds. Dynamic shear loading followed a sinusoidal waveform (0–10 N) at 1 Hz, simulating shear stresses observed during walking. These load conditions were designed to simulate physiologically relevant plantar stress levels reported during walking gait.

##### Baseline Session

Upon arrival at the gait laboratory, participants changed into standardised footwear (Sponarind 97308, Finn Comfort Inc. Hassfurt, Bavaria, Germany), thermally conductive socks (Silversock, Carnation Footcare, UK), sports tights (Run Tights, Karrimor Ltd., England), shorts and sport top (Sondico Core Shorts, Lovell Sports Ltd., England) which were worn throughout the entire experimental protocol. Participants’ height and weight were measured after they had changed into the standardised experimental clothing. They then sat on a chair and rested for 30 minutes with both feet elevated with the plantar surface offloaded to standardise baseline physiological conditions. A preliminary foot assessment was carried out, including visual inspection and participant temperature measurement. Temperature measurements were taken at the first metatarsal head, at the calcaneus and the forehead using an infrared thermometer (RS-8806S, RS PRO, London, UK). Participants in the group living with diabetes were assessed for loss of sensation in their feet using a monofilament test. Personal and clinical data—including age, foot dimensions, footwear preferences, physical activity levels, and foot health history—were collected using a structured assessment form. Ambient room temperature was measured at the beginning of the experimental protocol for each participant and remained constant with a mean of 21.4°c (± 1.3°c standard deviation). After 30 minutes, participant temperature and *PlantarSense* measurements were performed. *PlantarSense* measurements involved using the *PlantarSense* device to load the plantar tissue in a specific location (first metatarsal head [1stMTH] or calcaneus) with a loading profile in either compression or shear which included a preload (5N compressive load), slow ramped load up to a peak load (40N for compression or for shear, 10N in shear direction and 20N compression) followed by 10 cycles between the peak and preload at a frequency of 1Hz.

##### Walking Session

Participants then completed a 15-minute treadmill (Kingsmith WalkingPad, Beijing, China) walking trial at a self-selected pace. During walking, plantar loading data were recorded via F-scan pressure sensing insoles (Tekscan Inc., Norwood, MA, USA) which sampled data at a frequency of 50 Hz. Participant temperature and *PlantarSense* measurements were then performed again after the walking session finished.

##### Recovery Session

Participants then sat down and elevated their feet, so they were unloaded and remained in this position for 15 minutes to allow some recovery for the plantar tissue. Participant temperature and *PlantarSense* measurements were then repeated.

#### 2.2.3 Data Analysis and Statistics

##### Pressure time integral

Pressure–time integral (PTI) were extracted from in-shoe pressure data at defined regions matching the loading area of the *PlantarSense* device. The first metatarsal head region was located by placing the metatarsal head roughly in the centre of the ultrasound image from *PlantarSense* and then the probe was aligned in the anterior-posterior direction of the foot. A post-processing correction was performed from the pressure data using custom MATLAB code (The MathWorks Inc., Natick, MA, USA) to ensure that the peak pressure in that region measured from the Tekscan insole matched the position of the metatarsal head from the ultrasound image taken with *PlantarSense*. A similar procedure was used for the calcaneus region measurement. Pressures below 3 kPa were discarded to reduce noise. PTI was calculated from the pressure, P, measured in from every sensel of the Tekscan insole within the region of interest (from pixel index i=1 to N) multiplied by the time increment, *Δt*, over the total walking time (from t=0 to T), see Equation (3). The probe head (tissue loading area), A, was 33 mm × 9 mm and the loading time, T, was the full 15-minute walking duration.

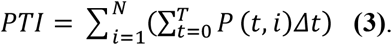

##### Statistical Methods

All data were anonymised prior to analysis to protect participant confidentiality. Given the present sample size (n = 30 feet), distributional assumptions were not uniformly satisfied across outcomes; analyses were therefore specified a priori on a variable-by-variable basis. Normality was assessed using the Shapiro–Wilk test within each group for every variable and comparison. For between-group contrasts (people living with diabetes vs controls), Welch’s two-sample t-test was used when both groups met normality; otherwise, the Mann–Whitney U test was applied. Within-group change was examined using change scores (Δ%), tested against zero: one-sample t-tests were used for approximately normal change scores, and Wilcoxon signed-rank tests were used when normality was not supported. Summary statistics are reported as mean ± standard deviation for normal distributed data and median (interquartile range) for non-normal distributed data.

Effect sizes of change and group differences were reported alongside p-values, following recommendations by Lakens [29]. For within-group analyses, Cohen’s dz was calculated as the mean change divided by the standard deviation of the paired differences, and interpreted as negligible (<0.2), small (0.2–0.5), medium (0.5–0.8) and large (>0.8). For between-group analyses, standardised differences were aligned with the inferential approach: Hedges’ g was used for parametric comparisons and interpreted as negligible (<0.2), small (0.2–0.5), medium (0.5–0.8) and large (>0.8); Cliff’s δ was used for non-parametric comparisons and interpreted as negligible (<0.147), small (0.147–0.33), medium (0.33–0.474) and large (>0.474). All analyses were performed in MATLAB (The MathWorks Inc., Natick, MA, USA). Tests were two-tailed with statistical significance set at p < 0.05, and significance was denoted as * p < 0.05, ** p < 0.01, and *** p < 0.001.

## Results

### 3.1 Bench top mechanical test validation

Accuracy from measurements on the silicone sample was 96% and 97% for the compressive and shear modulus respectively. Similarly high accuracy was observed for measurements on porcine tissue in vitro of 92% and 95% respectively for compressive and shear modulus. The *PlantarSense* System demonstrated high repeatability in modulus estimation, with intra-sample variability (SD/mean) <10% for both compressive and shear tests in silicone (Ecoflex™ 00-10, Smooth-On Inc., Macungie, USA) and porcine tissue.

### 3.2 Human test results

#### 3.2.1 Plantar pressure analysis during walking (pressure–time integral)

There was no statistically significant difference between groups (p = 0.898) for PTI in both the calcaneus and metatarsal head region due to the walking activity. At the first metatarsal head (1stMTH), the mean PTI was only 2.5% higher in diabetic group (456 MPa·s ±204 MPa) than control group (444 MPa·s ±257 MPa). The associated effect size was very small (g = 0.1), indicating negligible group separation at the forefoot. A similar pattern was observed at the calcaneus. Mean PTI at the heel was slightly higher in the diabetic group (1350 MPa·s ±314) than in the control group (mean = 1324.5 MPa·s ±288.1), but again this difference was not statistically significant (p = 0.836) and the effect size was negligible (g = 0.1).

#### 3.2.2 Planter tissue temperature

For temperature and temperature changes during activities, at the 1stMTH, both groups showed a similar post-walk temperature rise (Baseline→ After walk: diabetic group ΔT = 7.2% ± 7.3, control group ΔT = 6.7% ± 5.1, g = 0.1), and values remained slightly above baseline after recovery (Baseline → After recovery: diabetic group ΔT = 4.2% ± 5.1, control group ΔT = 2.1% ± 5.1), consistent with incomplete cooling (see figure 3.1a). At the calcaneus, walking produced a larger increase in the diabetic group (11.0% ± 4.7) than the control group (6.9 %± 3.7; p = 0.028) with a reported large effect size (g = 1.0), indicating a more pronounced activity-induced thermal response at the heel in diabetic group. The recovery changes were small but still above baseline (diabetic group ΔT = 2.3% ± 8.0; control group ΔT = 0.6 ± 4.2%, g = 0.3), suggesting residual warming particularly in the diabetic groups (see figure 3.1b). Core body temperature remained approximately unchanged (Δ≈0) across all sessions.

#### 3.2.3 Planter Tissue Energy Dissipation

Energy dissipation ratio (EDR) showed site- and direction-dependent patterns at baseline and with activity. In compression at baseline, EDR at the 1stMTH was significantly different between groups (diabetic group vs control group; p = 0.040, Fig. 3.3.a), with diabetic group showed a higher central tendency (mean of 67.8%, standard deviation 10.7% and mean of 56.0%, standard deviation 19.1%). By contrast, calcaneal compressive EDR did not differ significantly between groups at baseline with 8% lower in the diabetic group. Activity-related changes in compressive EDR (ΔEDR%) were present but highly variable, and between-group contrasts were non-significant at both anatomical sites. The larger changes were observed from 1stMTH region. Directionally, the ΔEDR suggested 23.7% reduction in the diabetic group and 22.7% reduction in the control group after walking, with recovery showing 27.1% reduction and 7.0% reduction respectively.

In shear, baseline EDR appeared to show weaker group separation than in compression (Fig. 3.3.d; Calcaneus: mean EDR in the diabetic group = 56.8% vs mean EDR in the control group = 52.4%). Activity-related ΔEDR in shear again demonstrated wide dispersion with no clear between-group differences at either site (Fig. 3.3.e–f), although within-group ΔEDR shifts in 1stMTH suggested significant difference showed in the control group (-24.5, %p=0.001 in Baseline → After Walking and -14.1%, p=0.009 in Baseline → After Recovery).

## Discussion

Hypothesis 1 stated that, plantar surface temperature change after activity and after recovery will be great and lower respectively in people living with than without diabetes. The observed thermal patterns support the hypothesis that people living with diabetes experience higher plantar heating and delayed cooling under mechanical load, however statistical significance was only confirmed in some cases.

Both the 1^st^ metatarsal head (1stMTH) and calcaneus regions exhibited statistically significant differences after walk temperature increases of 7.2% (1.7°C) and 11.0% (2.9°C) for the diabetic group and 6.7%(1.7°C) and 6.9%(1.8°C) for the control group. The difference between the diabetic and control group was only statistically significant for the calcaneus region after walking (p=0.03). During the recovery period, both the 1stMTH and calcaneus regions remained elevated above baseline, with residual temperature increases of 4.2% (1.0°C) and 2.3% (0.6°C), respectively, in the diabetic group, compared with 2.1% (0.5°C) and 0.6% (0.2°C) in the control group. Although these between-group difference did not reach statistical significance in 1stMTH, the effect size (g = 0.38) suggested a moderate tendency for higher residual temperature in the diabetic group at the 1stMTH, indicative of delayed thermal recovery at the forefoot. Core body temperature changes remained close to zero throughout all activities, indicating that the observed differences were local rather than systemic and thus unlikely to confound the plantar mechanical property measurements.

Serantoni et al. [33] conducted a 6-minute walk test showing thermoregulatory impairment in people living with diabetes, they used thermal imaging which revealed that those with diabetic peripheral neuropathy (DPN) failed to cool down as efficiently as those without, confirming a link between DPN and thermoregulatory dysfunction. These studies support the mechanism of microvascular and neural deficits leading to heat retention in the diabetic foot. In addition, mild inflammation or subclinical tissue injury may manifest as a local rise in plantar skin temperature; sustained focal “hot spots” have been widely reported as an early warning signal for impending diabetic foot ulceration and are used in preventive temperature-monitoring strategies. It should be noted that our study did not include people living with diabetes and peripheral neuropathy, hence, diabetes was likely less severe in our participants than in other thermal studies [27, 32, 33].

Regional differences in thermal response were observed between the heel (calcaneus) and forefoot (1stMTH), particularly in post-walk warming and recovery. The calcaneus in the diabetic group showed the greatest immediate temperature increase of all regions in both groups, indicating that the heel was the site most sensitive to walking-induced warming. In contrast, although post-walk temperature increases at the 1stMTH were not significantly different between groups, the 1stMTH in the diabetic group showed the smallest temperature reduction during recovery, indicating the least complete thermal recovery. These findings suggest impaired and region-specific thermoregulation in people living with diabetes, even in the absence of ulceration, characterised by greater heat accumulation at the heel and more persistent residual heating at the forefoot. Such delayed cooling may increase tissue vulnerability under repeated mechanical loading and supports the concept that impaired thermal regulation may act as a co-factor in ulceration when combined with pressure. This is an area for further research – e.g., whether interventions like cooling insoles or improving microcirculation (via medication or exercise) can normalize these thermal responses.

Our findings provide support for hypothesis 2 being partially true in that plantar tissue energy dissipation will be different in both compressive and shear in people living with than without diabetes and changes will be greater after activity and slower to recover in people living with than without diabetes. Statistical significance was only proved in some cases. The key finding are that the diabetic group showed a statistically significant higher EDR than the control group in 1stMTH in compression for baseline measurement. In shear at the 1stMTH, the control group demonstrated statistically significant reductions after walking (™24.5%, p = 0.001) with partial recovery after rest (™14.1%, p = 0.009), while diabetic group exhibited more variable, non-significant shifts. Following a short rest, controls tended to rebound toward baseline EDR, whereas diabetic group showed less complete restoration. For example, in 1stMTH compression, control group were only 7% below baseline after recovery, whereas the diabetic group remained 27% below baseline (and in some cases declined further from post-walk to recovery). A further important finding was that, despite only small absolute between-group differences in shear EDR, activity-related EDR responses were larger under shear than under compression. For example, at the calcaneus in the control group, post-walk EDR decreased by 5.4% in compression but by 21.5% in shear. This may indicate that plantar soft tissues are more responsive to recent loading exposure in the shear direction, which is notable given the recognised role of shear in diabetic foot tissue damage.

Under baseline conditions, group differences in EDR were strongly site- and direction-dependent, rather than uniform. In compression at the 1stMTH, the diabetic group exhibited a significantly higher EDR (mean =67.8%) than control group (mean= 56.0%; p = 0.040), indicating greater energy loss in the diabetic group. By contrast, baseline compressive EDR at the calcaneus was not significantly different between groups (49.8% and 54.3% in the diabetic and control groups respectively), and in shear at both sites the between-group differences were small (60.7% and 60.5% in 1stMTH, 56.7% and 52.4% in calcaneus for the diabetic and control groups respectively) and non-significant. Hysteresis was evident in specific conditions, most clearly the forefoot under compression, suggesting region-specific alterations in viscoelastic function.

This pattern is broadly consistent with previous evidence that plantar soft tissues in diabetic group can exhibit a more “lossy” mechanical response, while also showing marked site- and direction-specific heterogeneity. An ultrasound-indentation study reported significantly higher heel-pad EDR in type 2 diabetes than in controls (36.1% vs 27.9%, p < 0.001), suggesting greater impact energy dissipation that may contribute to tissue breakdown. Our baseline findings partly align with this, as elevated hysteresis was observed in the diabetic group at the first metatarsal head (1st MTH) under compression; however, the absence of a uniform effect across all sites and loading modes indicates that diabetes-related changes in EDR are not spatially homogeneous. EDR also tended to be higher at the 1st MTH than at the calcaneus, consistent with known regional differences in plantar tissue mechanics. This may reflect functional specialisation, with the forefoot adapted for load-bearing support and propulsion under combined compression–shear, whereas the heel pad is optimised for transient impact attenuation.

Mechanistically, elevated compressive EDR at the forefoot in the diabetic group is consistent with a more viscous and less elastically efficient response, in which a greater proportion of mechanical work is dissipated through internal friction rather than recovered elastically. Histological studies have reported collagen disorganisation and septal disruption or thickening in diabetic plantar fat pads, features associated with poor rebound and altered impact energy dissipation. A regional EDR gradient may nevertheless persist because the calcaneal and forefoot pads are structurally and functionally distinct: the calcaneal pad is thicker and more compartmentalised, favouring elastic support, whereas the thinner forefoot pad is exposed to greater multi-directional deformation and inter-compartmental sliding, increasing frictional losses. Consistent with this, diabetes may produce mixed mechanical signatures across regions rather than a uniform pattern, reflecting differences in local architecture, functional demands and in vivo boundary conditions.

Regarding activity effects, the second part of the hypothesis, greater post-walk increases in hysteresis and slower recovery in diabetic group, was not supported in the expected direction. Between-group differences in activity-induced ΔEDR were not statistically significant overall, but within-group effects and recovery patterns were informative. Immediately after walking, EDR decreased on average in both groups, at both sites, and in both loading modes. In compression at the 1stMTH and calcaneus, EDR significantly fell by 23.7%(p=0.02) and 4.3%(p=0.01) in the diabetic group and reduced by 22.7% and 5.4% in the control group, respectively, relative to baseline. In shear at the 1stMTH and calcaneus, EDR reduced by 27.1% and 6.7% in the diabetic group and reduced significantly by 24.5%(p=0.01) and 21.5%(p=0.01) in the control group, respectively. After recovery, EDR increased in both groups but remained below baseline on average across both sites and loading modes. In compression at the 1stMTH and calcaneus, EDR reduced by 27.1% and 8.0% in the diabetic group and reduced by 7.0% and 5.5% in the control group, respectively, relative to baseline. In shear at the 1stMTH and calcaneus, EDR reduced by 4.5% and 2.5% in the diabetic group and reduced by 14.1%(p=0.01) and 5.6% in the control group, respectively. Notably, statistically significant within-group reductions were observed in the diabetic group under compression at the 1stMTH both immediately after walking and after recovery, and in the control group under shear at the 1stMTH immediately after walking and after recovery, as well as at the calcaneus after walking. Together, these findings suggest that although activity-induced ΔEDR did not differ significantly between groups, loading produced measurable site- and mode-specific modulation of viscoelastic behaviour, with particularly clear responses at the 1stMTH and during shear recovery in the control group.

The counter-intuitive reduction in after walking results is compatible with viscoelastic “conditioning” or load-history effects: repeated cyclic loading can transiently reduce apparent dissipation by altering fluid distribution, pre-strain, and microstructural engagement, effectively changing the loop shape on subsequent cycles. The more clinically salient divergence emerged during recovery. Although between group significance was not demonstrated there was significance within individuals based on activity related changes—likely due to dispersion and limited power—these trends are consistent with faster restoration of viscoelastic function in controls and more persistent fatigue-like behaviour in people living with diabetes. This interpretation echoes Yang et al. [41], who reported that prior loading can reduce heel-pad thickness and viscous modulus in the short term, decreasing cushioning capacity. Even if both groups experience an acute reduction in damping following walking, a slower return toward baseline in diabetic group would extend the duration during which tissues operate in a mechanically compromised state.

Clinically, these hysteresis-related alterations may be relevant to tissue tolerance and injury risk in the diabetic group. Higher baseline hysteresis implies that more mechanical work is absorbed internally by the tissue each cycle, which can be protective in moderation but may also reflect inefficient elastic recoil and higher internal strain accumulation. Prior studies have linked elevated EDR to ulceration susceptibility. Hsu et al. [57] reported higher heel-pad EDR in people living with diabetes (particularly in those with ulcer history) and suggested that greater dissipated energy may increase ulcer risk. Our results extend this concern by suggesting that after activity, diabetic tissue may also recover more slowly, potentially prolonging periods of altered damping and load-sharing. In parallel, shear mechanics remain clinically important: even when shear EDR differences are modest, diabetes-related increases in shear stiffness can compromise the tissue’s ability to accommodate shear safely. Pai and Ledoux [42] reported substantially stiffer diabetic plantar tissues in shear and argued that this may compromise stress dissipation and increase ulceration risk. This is biomechanically plausible because shear deformation is strongly influenced by skin–subcutaneous coupling, septal architecture, and inter-compartmental sliding—all mechanisms that are readily perturbed by repetitive gait loading and are likely to recover more slowly when viscoelastic function is impaired. Taken together, the broader mechanical picture points to a reduced capacity to tolerate repetitive combined loading (compression + shear), particularly when recovery is incomplete. Over time, repeated walking bouts could therefore promote cumulative microdamage by exposing tissues to repeated stresses while still in a partially “fatigued” mechanical state, thereby increasing vulnerability to breakdown in the diabetic foot.

These results suggest that, following an equivalent bout of mechanical work (e.g. walking), plantar tissue in people living with diabetes not only exhibits a lower proportion of energy dissipation (i.e. reduced hysteresis), but also a more persistent post-activity mechanical state. In practical terms, tissue may remain relatively stiff and less compliant for longer after walking, plausibly reflecting slower restoration of interstitial fluid distribution and microstructural recovery. This provides a useful context for interpreting the accompanying thermal findings. Importantly, regarding the hypothesis 1, a lower EDR does not imply a cooler foot. Mechanical energy can be converted to heat through several pathways beyond bulk tissue hysteresis, including friction at the skin–footwear interface, internal shear at tissue boundaries, and muscular effort associated with altered gait mechanics [18,39,40]. Moreover, heat removal is primarily governed by thermoregulatory capacity—cutaneous perfusion and sweating—rather than by hysteresis alone. In healthy individuals, walking typically elicits vasodilation and sweating, enhancing convective/radiative heat loss and evaporative cooling. In people living with diabetes, microvascular dysfunction and sweat gland impairment can blunt these responses, promoting heat retention even when tissue-level hysteresis is lower. In addition, stiffer, less deformable tissues may concentrate stresses, predisposing to reactive hyperaemia or low-grade microtrauma that can further elevate local temperature after activity. Overall, the balance between heat generation and heat dissipation is likely shifted towards retention in the diabetic foot.

In summary, diabetes did not produce a uniform increase in plantar hysteresis across sites and directions. Instead, EDR differences were context-specific, with baseline elevation most evident at the 1st metatarsal head under compression and trend-level evidence of less complete post-walk restoration in people living with diabetes, consistent with impaired recovery of viscoelastic function after everyday loading. Clinically, the forefoot and shear direction appears to be the most concerning region because it combines altered energy dissipation with greater thermal burden, a pattern that is consistent with the predominance of forefoot “hotspots” and ulceration relative to the heel and likely reflects site-specific differences in load sharing, tissue architecture and perfusion. These findings suggest that plantar thermal responses should be interpreted alongside mechanical metrics: an apparently acceptable pressure profile may still coexist with elevated post-walk temperature, potentially signalling subclinical microvascular impairment and insufficient mechanical buffering. Together, this supports prevention strategies that address both sides of the problem—reducing repetitive combined compression–shear exposure and improving load attenuation (e.g., cushioned insoles, custom orthoses and activity dosing) while also considering interventions that support thermal clearance and vascular function—thereby limiting cumulative tissue stress (through activity pacing) and mitigating diabetes-related vulnerability after walking.

A novel measurement device, *PlantarSense*, was designed and used for measuring in vivo plantar tissue mechanical properties under both compressive and shear loading across dynamic conditions. Bench-top validation using silicone and porcine samples showed an accuracy and repeatability greater than 92% and 90% respectively.

The groups living with and without diabetes were selected for the study and had very similar PTI for walking activity (less than 2% difference) and similar ages. Under the present experimental conditions, diabetic and control group experienced very similar cumulative plantar loading at both the 1stMTH and calcaneus during walking. This contrasts with some previous studies that reported substantially elevated plantar pressures or PTI in individuals with diabetes, particularly in the presence of peripheral neuropathy or established foot deformity, but those studies were often limited by confounding factors such as higher body mass index, altered gait patterns, or inclusion of patients with a history of ulceration[41,42]. Our groups were age, BMI, sex matched within 4.2%, 5.5%, 0 % respectively. Whereas other studies either didn’t report this or used people living with diabetes who were more severely affected, had higher BMI and had altered gait through peripheral neuropathy by their condition. Consequently, the observed activity-related changes in plantar tissue mechanics are unlikely to be driven by differences in cumulative external loading, and more plausibly reflect intrinsic tissue behaviour and recovery capacity. However, a limitation is that our diabetic group cohort may have been relatively high-functioning, potentially underestimating loading-related disparities and biomechanical impairment seen in more severely affected populations.

Several limitations should be acknowledged. A small sample size meant analyses were underpowered and many between-group comparisons were non-significant (p > 0.05); findings should therefore be considered as pilot study. Recovery was assessed at a single post-rest time point and the walking task represented only one loading “dose”, limiting inference about recovery kinetics and dose–response effects; residual variability in gait strategy may also have contributed to dispersion in activity-induced changes. Finally, for ethical and injury risk reasons the maximum indentation force was capped at 40 N (≈135 kPa) to avoid discomfort; this loading range may not fully represent peak gait pressures and could influence estimates of nonlinearity, hysteresis and functional relevance under real-world walking loads.

Although exploratory, the effect directions and variability in temperature and EDR support the need for adequately powered follow-up studies (e.g. with n≥29 per group, larger if stratifying by diabetes complications) using mixed-effects models to test diabetes severity, foot region, loading direction and activity pacing while adjusting for key confounders (age, BMI, foot structure, gait speed and recovery timing/duration). Studies should integrate imaging and/or multi-layer modelling to resolve layer/compartment heterogeneity, sample recovery at multiple post-activity time points and graded walking “doses”. Also, where ethically feasible extending the mechanical loading range via safe, incremental protocols to better approximate peak gait plantar stresses.

## Conclusion

This study developed and validated *PlantarSense*, a novel ultrasound-loadcell integrated device for quantifying in vivo plantar soft-tissue mechanics under dynamic compressive and shear loading. Using this device, we identified significant activity-related changes in both plantar temperature and energy dissipation ratio (EDR), indicating that recent loading modulates both the thermal and viscoelastic behaviour of plantar tissues, which differed in people living with and without diabetes. People living with diabetes exhibited greater post-walk temperature increases than controls, most notably at the calcaneus, where the increase was significantly greater (11.0% vs 6.9% in the control group). During recovery, residual temperature elevation remained greater in people living with diabetes, particularly at the first metatarsal head (4.2% vs 2.1% in controls), suggesting less complete thermal recovery and impaired thermoregulatory capacity.

In parallel, EDR decreased after walking in both people living with and without diabetes, but the magnitude and recovery of this response were foot region and loading mode dependent. Baseline compressive EDR at the 1st metatarsal head was significantly higher in people living with diabetes (67.8%) than without (56.0%), while activity-related EDR modulation was generally greater under shear (21.5%) than under compression (5.4%) for participants without diabetes, suggesting that plantar tissues may be especially sensitive to shear-related loading history and recovery. Following a short rest, controls tended to recover towards baseline EDR, whereas the diabetes group showed less complete restoration, indicating a reduced capacity to regulate viscoelastic behaviour in response to activity.

Taken together, these findings suggest that dynamic thermal and mechanical assessment have the potential to reveal aspects of plantar tissue dysfunction that are not captured by static measures alone. Combined evaluation of temperature and EDR may therefore offer a more sensitive approach to characterising impaired plantar tissue adaptation and recovery in people living with diabetes, with potential to improve mechanistic understanding of diabetic foot vulnerability and support future ulcer risk stratification. Future work should include larger, adequately powered studies to confirm these findings and determine their clinical utility. In addition, the development of a more controlled loading system capable of applying larger but safe physiological loads would allow more representative simulation of gait-related stresses and improve assessment of plantar tissue behaviour under clinically relevant conditions.

## Data Availability

All data produced in the present study are available upon reasonable request to the authors

